# Diffusion Signals of Glial Activation Correlate with Fitness Scores in Parkinson’s Subcortical Gray Matter

**DOI:** 10.1101/2024.11.27.24318085

**Authors:** Benjamin T. Newman, Ryan Foreman, Erin Donahue, Michael Jakowec, Andrew J. Petkus, Daniel P. Holschneider, Joseph O’Neil, Dawn M. Schiehser, Giselle Petzinger, John Darrell Van Horn

## Abstract

The cholinergic basal forebrain and subcortical gray matter have previously been implicated in Parkinson’s disease (PD) but the relationship between the symptomology of PD, including cognitive impairment, and specific patterns of damage remains unclear. Additionally understanding the impact of fitness, including motor skill fitness, on brain structure remains a gap in knowledge. In this study, we examine a longitudinal cohort of PD patients using advanced microstructural analysis derived from diffusion MRI, alongside volumetric, and connectivity components. These imaging metrics are compared to comprehensive cognitive and motor skill fitness metrics. Though the volumetric components well characterize change across the longitudinal timescale, the microstructural analysis was related to a number of composite cognitive scores including attention, executive function, cognition, language, as well as the level of motor skill fitness. We further assess the connectivity of the cholinergic basal forebrain, particularly the nucleus basalis of Meynert in area 4 (CH4), and find that connectivity between CH4 and the thalamus is associated with latent motor skill fitness. The pattern of microstructural results suggests that increased cellularity, potentially from astrocytic glial cells, may support reduced cognitive decline in individuals with PD as well as higher motor skill fitness.

## Introduction

Alongside its well-known motor symptoms, wide-ranging cognitive deficits are common in Parkinson’s disease (PD) ^1,2^. Per the Braaks’ staging hypothesis^3^, PD develops gradually as a neurotoxic aggregate form of alpha synuclein protein accumulates in structure after structure of the brain roughly in stages. As alpha synuclein load rises and neurons perish in each region, symptoms emerge reflecting interdiction of the normal functions of the region and/or its efferent targets. Thus, the hallmark motor symptoms of PD stem primarily from loss of pigmented neurons in the substantia nigra pars compacta and consequent interruption of their dopamine (DA) projections to the caudate and putamen^4^. Cognitive symptoms, the focus of this report, in contrast, have been partly attributed to synuclein pathology in the Ch4 portion of the nucleus basalis of Meynert in the substantia innominata and to interruption of their acetylcholine (ACh) supply to the amygdala and neorcortex^5–7^. Other segments of the cholinergic basal forebrain, including the medial septum (Ch1) and vertical (Ch2) and horizontal (Ch3) limbs of the diagonal band of Broca and their main target the hippocampus, are similarly afflicted^3^. (We refer below to the closely adjacent Ch1, Ch2 and Ch3 cholinergic centers collectively as “Ch123”.) The Braaks’ hypothesis implies that appropriate *in vivo* non-invasive neuroimaging of brain structures compromised by PD may aid clinicians and researchers in diagnosing and staging the disorder, making prognoses, identifying the anatomic sources of symptoms, and perhaps even designing novel therapies and predicting treatment outcomes.

These goals have been pursued intensively using magnetic resonance (MR) methods. Thereby, conventional morphometric structural MRI may prove too coarse to image the small nuclei implicated in PD, such as Ch4 and Ch123, quantifiably and reliably^8,9^. Diffusion microstructure imaging, in contrast, affords higher resolution and greater detail than macroscopic volumetric measures in characterizing symptom-generating structural degradation in PD^10^. Microstructural metrics can sample inside intra-axonal spaces, detect activated microglia and astrocytes, and measure changes in extracellular water content, such as those left by neuronal atrophy^11^. A great deal of work has been focused on diffusion tensor imaging-based measures of microstructure due to the ease of calculation. For example a prior study described widespread changes in fractional anisotropy (FA) using tract based spatial statistics in PD patients with psychosis compared to PD patients without psychosis^12^. Another study similarly found changes in FA associated with cognitive and executive function deficits with these associations being located in different segments of WM tracts depending on the cognitive process being assessed, further demonstrating that different symptom profiles may arise from different neurological patterns of damage^13^. Microstructural differences in FA have been observed between PD patients with cognitive impairment and those without, even in the absence of other macrostructural, cortical thickness, or volumetric differences^14^. Other studies using more advanced microstructural models have sought to more closely linked imaging metrics with underlying cellular changes. For example, studies have described increased extracellular free water in the substantia nigra in PD patients, indicating neuroinflammation or atrophy^11,15^.

This detail provided by microstructural analysis is important, not only to well characterize neuronal structure in PD, but also to explore the involvement of additional cellular types, which may represent new treatment targets. A series of studies in mouse models has suggested that glial markers, particularly astrocytic markers, are active in PD^16,17^. The above-mentioned capacity of diffusion microstructural methods to probe not only neuronal, but also astrocyte, status may be of particular relevance to PD. Our preclinical work indicates that morphological changes in astrocytes may be related to exercise and fitness and that activation of astrocytes may underlie the beneficial effects of exercise in PD^16,17^ Looking for specific regions of the brain that might be vulnerable to change early in PD symptomology, the cholinergic basal forebrain, and the nucleus basalis of Meynert in area 4 (CH4) has been implicated in a number of studies as involved in the spread of PD and in specific PD symptomology^5–7^.

This study utilizes several advanced diffusion microstructure metrics to characterize effects of PD at the cellular and sub-cellular level. These metrics (together with conventional structural MR volumetry) were sampled bilaterally in nine subcortical structures: the amygdala and hippocampus; the caudate, putamen and nucleus accumbens; the thalamus and globus pallidus; and Ch4 and Ch123. These structures vary in the degree they suffer from PD pathology and in their relationship to the central DAergic and cholinergic systems attacked by PD. Among these structures, the amygdala and hippocampus are major mesial temporal lobe (MTL) regions. Some amygdalar subnuclei already evince alpha-synuclein load at Braak Stage 4; the remaining subnuclei and the entire hippocampus are impacted by Stages 5-6^3^. Neither the amygdala nor the hippocampus is appreciably recipient of DAergic innervation from the substantia nigra pars compacta. The amygdala (alongside the neocortex) is innervated with ACh by Ch4 and the hippocampus by Ch123^18,19^. The caudate, putamen, and nucleus accumbens (ventral striatum) are the three nuclei composing the human striatum. None is a direct site of synuclein accumulation in PD. The caudate and putamen, however, are indirectly but strongly impacted by PD in that they are the major sites of DA output from the compacta^20^. The accumbens, in contrast, receives its DA input form the ventral tegmental area (VTA)^21^. Striatal nuclei receive only minor ACh input from midbrain or forebrain centers as their ACh innervation is primarily intrinsic, from cholinergic interneurons^22^. The globus pallidus is not a major site of synuclein accumulation in PD and likely recieves only secondary DAergic input from the compacta^23,24^. Nonetheless, the globus pallidus (especially its internal segment) is highly relevant to the motor symptoms of PD and their treatment as it is a node in both the direct and indirect pathways^25^. The globus pallidus probably receives it ACh input from the pedunculopontine (Ch5) and the laterodorsal tegmental nuclei (Ch6)^22^. Some subnuclei of the thalamus are burdened by synuclein in Stage 4^3^. The thalamus does not receive appreciable DA input from the compacta. The major cholinergic input to the thalamus is from Ch5-Ch6^26^. Ch123 suffers synuclein deposition in Stage 3, Ch4 in Stages 3-4^3,27–30^. Neither basal forebrain structure receives substantial DA input from the compacta^31^. As indicated, both serve as major sources of cholinergic output to the rest of the brain^32,33^. Using diffusion tractography, we additionally evaluated the structural connectivity between each structure and Ch4.

The microstructural metrics were acquired longitudinally in a cohort of PD patients with well-characterized motor and cognitive symptoms. We assessed the relationships between each regional MR metric and symptom change between baseline and follow-up. Though exercise was not a planned intervention in this sample, a “latent fitness” measure was acquired at baseline and 18-month follow-up in each patient. This permitted us to assess effects of fitness on MR endpoints.

## Methods

### Participants

Subjects diagnosed with PD were recruited from a cohort who were participating in an observational study examining associations between motor skill fitness and cognitive function over a two-year period. Participants were recruited through movement disorder clinics at both the Departments of Neurology, University of Southern California and the Veterans Affairs San Diego Healthcare System/University of California, San Diego. After performing quality control on the imaging data and excluding any subject with missing data, 22 subjects (12 female) was used in the final analysis. Subject mean age at baseline was 63.83 years ± 9.70 SD (male 69.29 years ±7.93 SD; female 60.36 years ±6.37 SD). Subjects had been diagnosed with PD on average 3.72 years ±2.65 SD (male 3.14 years ±1.74 SD, female 4.09 years ±3.04 SD) prior to study baseline. Subjects returned for follow-up evaluation at approximately 24 months. All imaging and clinical assessments were obtained at baseline and at approximately 24 months follow up. All measurements were obtained in the ‘ON’ medication state. Informed consent was obtained from all participants and all study procedures were approved by appropriate Institutional Review Board (IRB). A summary of all participants metrics in cognitive and fitness tests are reported in Table 1.

**Table 1:** Patient demographics and motor and cognitive symptoms of PD at baseline and 18-month follow-up. Also listed are baseline and follow-up latent fitness scores. Physical exercise is a well-known recently proposed therapy for PD. Although patients were neither encouraged nor discouraged from exercising as part of the study, many PD patients will adopt increased exercise on their own. Thus, this fitness metric was assayed to assess potential effects of fitness on study outcomes. Data are numbers of patients and/or mean ± standard deviation across the entire sample of N=22. P-values are for linear mixed model F-tests comparing follow-up to baseline scores controlling for age, sex, years after PD diagnosis, total brain volume, days between the baseline and follow-up visit, and scanning site. MDS-UPDRS=Movement Disorders Society-Unified Parkinson’s Disease Rating Scale Motor Score Part III, standard clinical test of motor symptoms of PD; high scores are “bad”. MOCA=Montréal Cognitive Assessment, test of global cognition commonly used as a clinical screener for dementia; high scores are “good”. Z-Scores were computed as composites of multiple tests in each of four cognitive domains (see text); high Z-Scores are “good”. Latent Fitness is an in-house developed measure of physical fitness with higher scores representing superior performance.

| Measure | | | Units | Baseline | Follow-Up | $p$-longitudinal |
| --- | --- | --- | --- | --- | --- | --- |
| *Demographics* |  |  |  |  |  |  |
| **Sex** |  |  | #m/#fm | 10/12 | -- | -- |
| **Age** | | | years | 63.8 ± 9.7 | -- | -- |
| **Formal Education** | | | years | 17.27±1.6 | -- | -- |
| **Time Since First Diagnosis** | | | years | 3.7 ± 2.6 | -- | -- |
| *Motor Symptoms* |  |  |  |  |  |  |
| **MDS-UPDRS** | | | Total Score | 20.89±9.71 | 15.50±20.11 | 0.1538 |
| *Cognitive Symptoms* |  |  |  |  |  |  |
| **MOCA** | | | Total Score | 26.57±2.27 | 26.30±2.52 | 0.6799 |
| **Executive Function** | | | Z-Score | 0.174±1.01 | -0.085±1.37 | 0.4088 |
| **Attention** | | | Z-Score | -0.00075±0.988 | -0.188±1.27 | 0.5262 |
| **Language** | | | Z-Score | 0.227±1.00 | 0.1633±1.02 | 0.807 |
| **Memory & Visuospatial** | | | Z-Score | 0.201±0.772 | 0.025±0.930 | 0.4285 |
| *Motor Performance* |  |  |  |  |  |  |
| **Latent Fitness** | | | Total Score | 0.235±0.368 | 0.352±0.423 | 0.4249 |

### Measures of Cognitive Performance

The Montreal Cognitive Assessment (MOCA)^34^ was administered as screening of general cognitive ability. The MOCA is a screening measure of cognitive impairment in people with PD recommended by the MDS^35^. Participants completed tests of attention, working memory, language, visuospatial function, executive function, and episodic memory.^36^.Attention was measured by (i) the number of seconds to complete the color naming subtest from the Delis Kaplan Executive Function System (D-KEFS) Color Word Interference Test (CWIT)^37^, (ii) the number of seconds to complete the word reading subtest from the CWIT, and (iii) the total score for the forward digit span condition of the Adaptive Digit Ordering Test (DOT-A)^38^. The total raw score from the digit sequencing condition of the DOT-A was used to measure working memory. The confrontational naming aspect of language ability was measured by the total number of correct responses provided spontaneously or with semantic cueing on the Boston Naming Test^39^. The verbal fluency aspect of language was measured by the total correct number of words on the D-KEFS Letter Fluency test (phonemic fluency), while the D-KEFS Category Fluency test measured semantic fluency. Verbal episodic memory was measured by the total number of correct words recalled across the five immediate recall trials (IR) of the 2nd Edition of the California Verbal Learning Test (CVLT-II)^40^ and the total number of words recalled on the long-delayed free recall condition of the CVLT-II (CVLT-II LD). Non-verbal episodic memory was measured by immediate and delayed recall from the Brief Visuospatial Memory Test-Revised^41^. Visuospatial ability was measured by the total raw score on the Judgment of Line Orientation Test^42^ and the total raw score on the Hooper Visual Organization Test^43^. Executive function was measured using the number of perseverative responses on the Wisconsin Card Sorting Test-64^44^, the number of seconds to complete the color-word inhibition subtest of the D-KEFS CWIT, the number of seconds to complete the inhibition/switching subtest from the D-KEFS CWIT, and the number of correct words produced on the verbal fluency switching subtest from the D-KEFS verbal fluency test. These cognitive evaluations were then z-scored and collated into a single metric for each individual cognitive domain, including attention, working memory, language, visuospatial function, executive function, and episodic memory.

Participants self-reported their age, years of formal education, and sex and years of diagnosis. The Montreal Cognitive Assessment (MOCA)^34^ was administered as screening of general cognitive ability. The MOCA is a screening measure of cognitive impairment in people with PD recommended by the MDS^35^. Participants self-reported their age, years of formal education, and sex. Motor symptom severity was assessed using the Movement Disorders Society-Unified Parkinson’s Disease Rating Scale (MDS-UPDRS) motor score part III^45^.

### Measurement of Motor Skill Fitness

In recent years, potential beneficial effects of physical exercise on PD, especially its cognitive symptoms, have been investigated intensively^46^, including by our laboratory^16,17^. The effects of exercise on PD were not part of the original specific aims of this study and patients were neither encouraged nor discouraged from changing their exercise habits between baseline and follow-up. Knowledge of the benefits of exercise, both generally and specifically for PD, is, however, widespread in clinical populations and many PD patients are prone to increase their level of physical exercise on their own, even without a physician’s recommendation. We felt it would be remiss not to take this factor into account. Therefore, as an adaptive strategy, each patient underwent an assessment of physical fitness yielding a Latent Fitness Score both at baseline and at follow-up. Motor skill fitness was assessed using objective and standardized clinical measures that include upper limb fine and coarse motor functions, balance, coordination, agility, and endurance, and that requires timed performance in most items. Objective assessments included (1) the Physical Performance Test (PPT)^47^, (2) the Short Physical Performance Battery (SPPB)^48^, (3) the Timed Up and Go (TUG) test^49^, and (4) the Ten-step Test (TST)^50^. For this study a latent factor analysis approach was used to estimate motor skill fitness at baseline and follow up for each study participant. The advantage of the latent factor approach is (i) it captures the variance shared among multiple individual tests within a construct (e.g. motor skill fitness) and ultimately minimizing measurement error, (ii) it reduces the number of statistical tests and (iii) provides a means to aggregate data that represent a similar theme or construct. First to determine the latent factor for motor skill fitness, raw scores for all motor skill assessments, including the PPT, SPPB, TUG and TST were transformed so that for all tests higher scores represented better performance. Standardized z-score for each test was then calculated based on the mean and standard deviation. Next, a latent factor for motor skill fitness was generated using a structural equation modeling (SEM) that loaded all four clinical assessments. The SEM estimating the motor skill fitness latent factor had strong model fit (Chi-square = .021 (2) p = .90; Confirmatory Fit Index = 1.00; Root Mean Square Error of Approximation = <.001).

### Image Acquisition

Diffusion, T1-weighted, and T2-weighted images were acquired from each subject at baseline and follow-up. Diffusion images were acquired with an isotropic voxel size of 1.7×1.7×1.7mm^3^, 120 non-colinear gradient directions at b=3000 s/mm^2^, 30 at b=1300 s/mm^2^, 30 at b=300 s/mm^2^, 12 at b=80 s/mm^2^, and 12 b=0. T1-weighted MPRAGE images were collected with a FOV of 208×300×320, and an isotropic voxel size of 0.8×0.8×0.8mm^3^. Images were acquired on a Siemens 3T Prisma system at USC and on a GE Healthcare 3T MR 750 system at the San Diego VA. Acquisition was identical at each site with the exception of the MPRAGE having a TR/TE=2400/2.22 ms at USC and TR/TE=2852/3.548 ms at San Diego. Recruitment site was included as a factor in all imaging analysis.

### Structural MRI Post-Processing

All images were analyzed in line with previously published research^51–53^. In brief, T1w images were analyzed using the *recon-all* pipeline in Freesurfer for volumetric components^54^. Freesurfer automatically generated gray matter masks (regions-of-interest—ROIs) for multiple subcortical structures taken from the Destrieux cortical atlas^55^ bilaterally and calculated the volume of each. All microstructural metrics below were averaged within these ROIs. MR endpoints were averaged across left and right of each ROI. ROIs included the amygdala and hippocampus; caudate, putamen, and nucleus accumbens; globus pallidus and thalamus.

### Diffusion MRI Post-Processing

Diffusion images were denoised ^56^, corrected for Gibbs ringing artifacts ^57^, and corrected for inhomogeneity fields using FSL’s *topup* and *eddy* commands utilizing outlier detection and replacement ^58–60^, The final preprocessed diffusion images were up-sampled to an isotropic voxel size of 1.3×1.3×1.3mm^3^ ^61^ and only the b=3000 s/mm^2^ images used as an outer b-value shell. White matter (WM), GM, and cerebrospinal fluid (CSF) tissue response functions were generated using the Dhollander algorithm^62^, and single-shell 3-tissue-constrained spherical deconvolution were used to generate the WM fiber orientation distribution (FODs) and GM and CSF representations. 3-Tissue Constrained Spherical Deconvolution^63–66^ was used to calculate the voxel-wise maps of the fraction of signal arising from each of 3 compartments: an intracellular anisotropic (ICA), intracellular isotropic (ICI), and extracellular isotropic freely diffusing water (ECI) compartment by setting the sum of all FOD coefficients equal to unity. WM-FODs were then used to create a cohort-specific template with a subset of all individuals in the cohort^67^. All subject’s WM-FODs were registered to this template using an affine non-linear transform warp with reorientation of the FODs, and then the template was registered to a b-value matched template in stereotaxic MNI space^68,69^. A fixel-based morphometry (FBM)^67,70^ approach was used to estimate the intra-axonal cross-sectional area within each voxel to be used as an apparent axonal volume fraction (AVF). A T2w image was obtained by extracting and averaging the b=0 image from the preprocessed diffusion images. T1w and T2w images were processed as described in the MICA-MNI pipeline^71^, including N4-bias correction^72^, rescaling both images from 0-100, co-registration using a rigid transform^72^, and subsequently registration to the cohort specific template using the same warp as the diffusion images. The aggregate g-ratio was calculated on a voxel-wise basis according to Stikov et al. and was used according to Mohammadi & Callaghan, identically to the calculation in Newman et al^52,73–76^. As a measure of intra-axonal volume, the fiber density cross section was used as the AVF ^67^. As a metric of myelin density, the T1w/T2w ratio was used as the myelin volume fraction (MVF). Aggregate conduction velocity was calculated based on the calculations of Rushton ^77^ and Berman et al. ^78^, again, identically to the calculation in Newman et al^52^. Each of these 6 different voxel-wise microstructure metrics are displayed in Figure 1, and were calculated for each voxel of the brain, from each subject, at both baseline and follow-up for this study.

**Figure 1:**
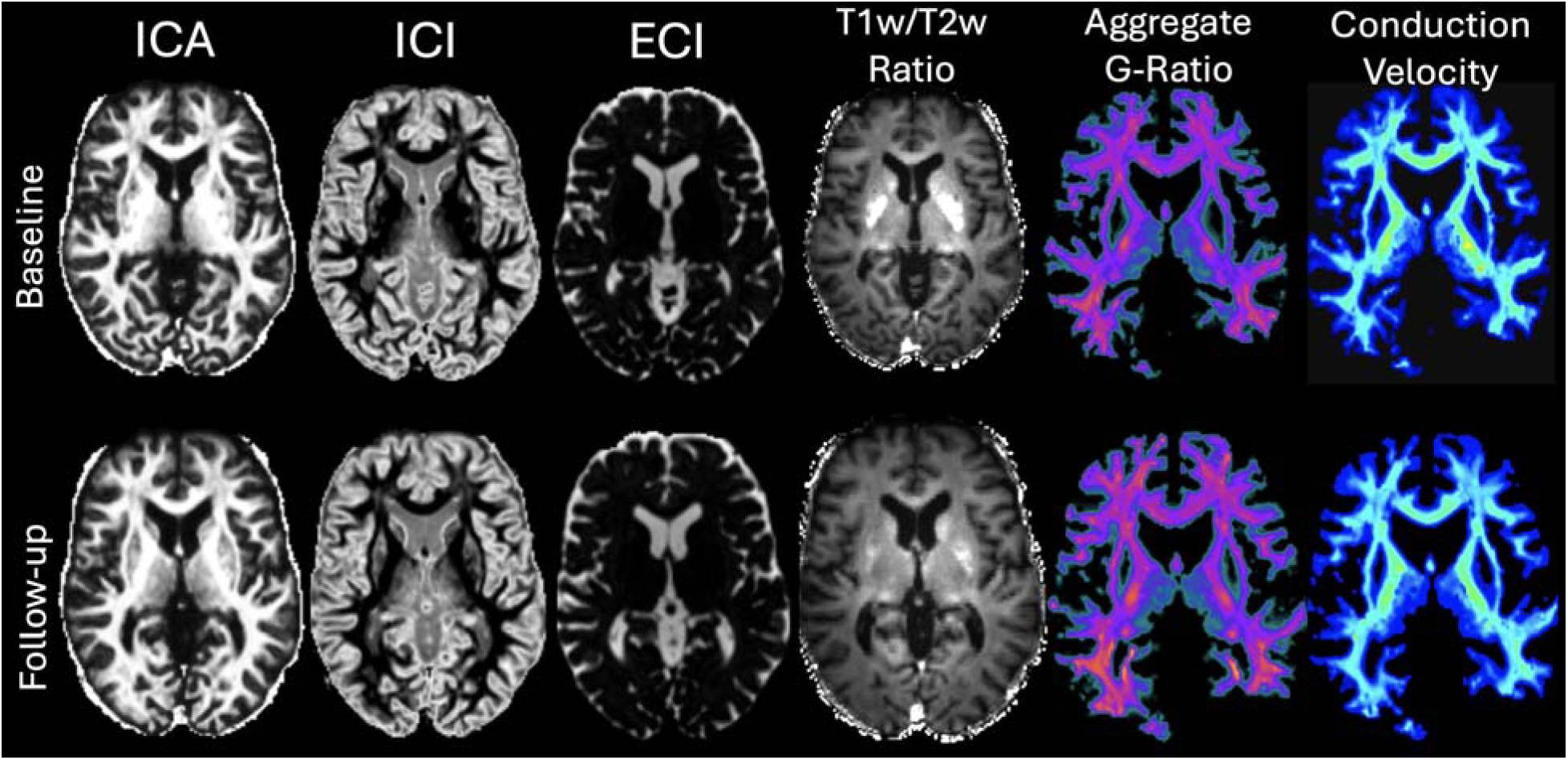
Representative native-space maps of each microstructural metric analyzed in this study from baseline and follow-up scans from a single subject. Each microstructural metric constitutes a single voxel-wise scalar value for mean aggregation within an ROI.

### Diffusion Tractography

Whole brain tractograms were generated for each subject at baseline and follow-up using Mrtrix3^79^. Diffusion image WM-FODs generated previously were used to derive principal direction of diffusion in each brain voxel, and the output from Freesurfer was used to generate a white matter – gray matter interface mask for seeding for an anatomically constrained tractography approach. Ten million tracts were generated using the iFOD2 probabilistic tracking algorithm which was then reduced to 2 million based on the FOD lobe integral using the spherically informed filtering of tracts (SIFT) algorithm^79,80^.

SIFT removes tracts in a systemic manner reliant on the underlying axonal anatomy to ensure remaining tracts are related to connectivity strength and allowing for direct comparison between subjects.

The goal of tractography was to quantify the diffusion-based structural connectivity between four of our subcortical ROIs and Ch4 (the location of which is displayed in Fig. 2). A tract rendered by the algorithm as described above was counted as connecting Ch4 with the ROI and included in the count for statistical analysis if the tract crossed both Ch4 and the ROI in question. The “structural connectivity” of each Ch4-ROI pair was equal to the number of tracts in the count. The four ROIs (amygdala, caudate, putamen, thalamus) were selected based on known Ch4 afferents from histological examination that overlapped with ROIs included in this study^81^.

**Figure 2:**
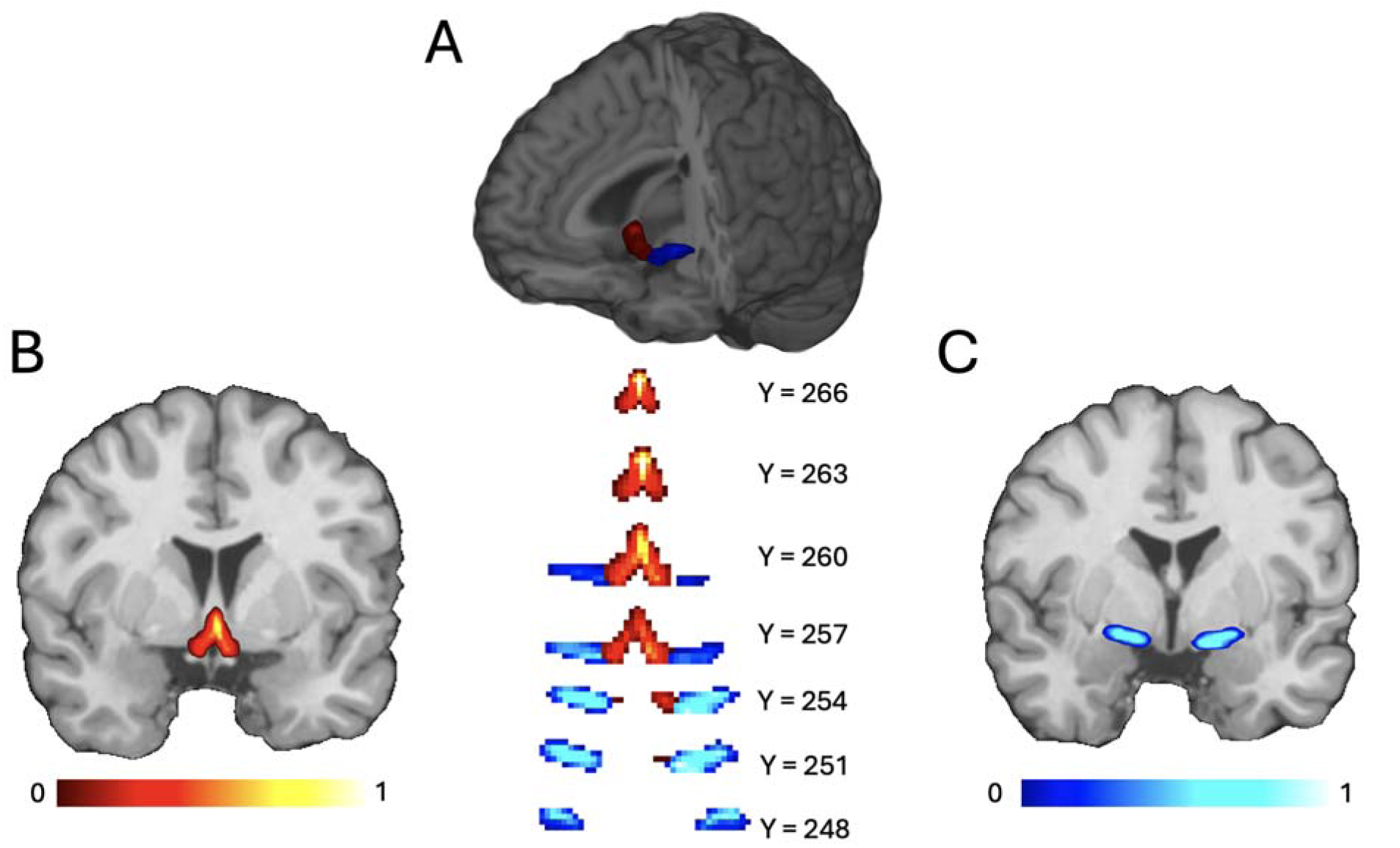
(A) Illustration of the placement of the Lorio et al. (B) CH4 and (C) CH123 probabilistic masks. The masks were thresholded at 80% probability to reduce noise from anatomical or registration placement.

### Statistical Analysis

All analyses were made using the linear mixed effects (*lmer*) package in R using type III F tests. All models were controlled for using subject age, sex, years after PD diagnosis, total brain volume without ventricles calculated by Freesurfer, days between the baseline and follow-up visit, and scanning site. These confounding variables were selected due to scientific merit and previously observed interactions with microstructural measurements^53,82,83^.

The first analysis examined longitudinal changes from baseline to follow-up in motor symptoms, cognitive symptoms, and latent fitness. There was no correction for multiple comparisons for the first analysis. The second analysis examined the same in volume and the six microstructural metrics in the nine ROIs. The third analysis examined the effects of baseline values of volume and each of the six microstructural metrics in each ROI on longitudinal changes in motor symptoms, cognitive symptoms, and latent fitness. Effects on latent fitness were also assessed using both baseline and follow-up latent fitness scores separately. The fourth analysis examined longitudinal cha ges in structural connectivity for each of the four Ch4-ROI connectivity pairs. The fifth analysis examined the effects of baseline values of structural connectivity for each of the four Ch4-ROI pairs on longitudinal changes in motor symptoms, cognitive symptoms, and latent fitness and separately on baseline and follow-up fitness. For the second and subsequent analyses multiple comparisons were corrected for using false discovery rate (FDR) under the Benjamini-Hochberg procedure.

## Results

### Imaging metrics

Summary values for each microstructural metric in each ROI can be found in Figure 3. These values and standard deviations are numerically presented for each timepoint in Supplementary Table 1. There was no significant difference in any microstructural metrics after Benjamini & Hochberg analysis between baseline and follow-up. The summary values for each of the volumetric subcortical ROIs available from Freesurfer are presented in Figure 4. There was a significant difference between baseline and follow-up in the hippocampus (F=16.65, p<0.001), pallidum (F=11.22, p<0.001), putamen (F=57.61, p<0.001), caudate (F=129.25, p<0.001), and thalamus (F=13.21, p<0.001). There was no significant difference between the baseline and follow-up volume of the accumbens (F=1.56, p=0.211 n.s.). Interestingly there was no significant relationship between scan time and total brain volume in the models considering the putamen, hippocampus, or pallidum, suggesting that these areas may be changing at similar rates to the brain as a whole, while in the caudate, and thalamus both scan and total brain volume were highly significant, suggesting that these areas may be changing differentially than total brain volume, by gaining relative volume, rather than being reduced in volume (Fig. 4).

**Figure 3:**
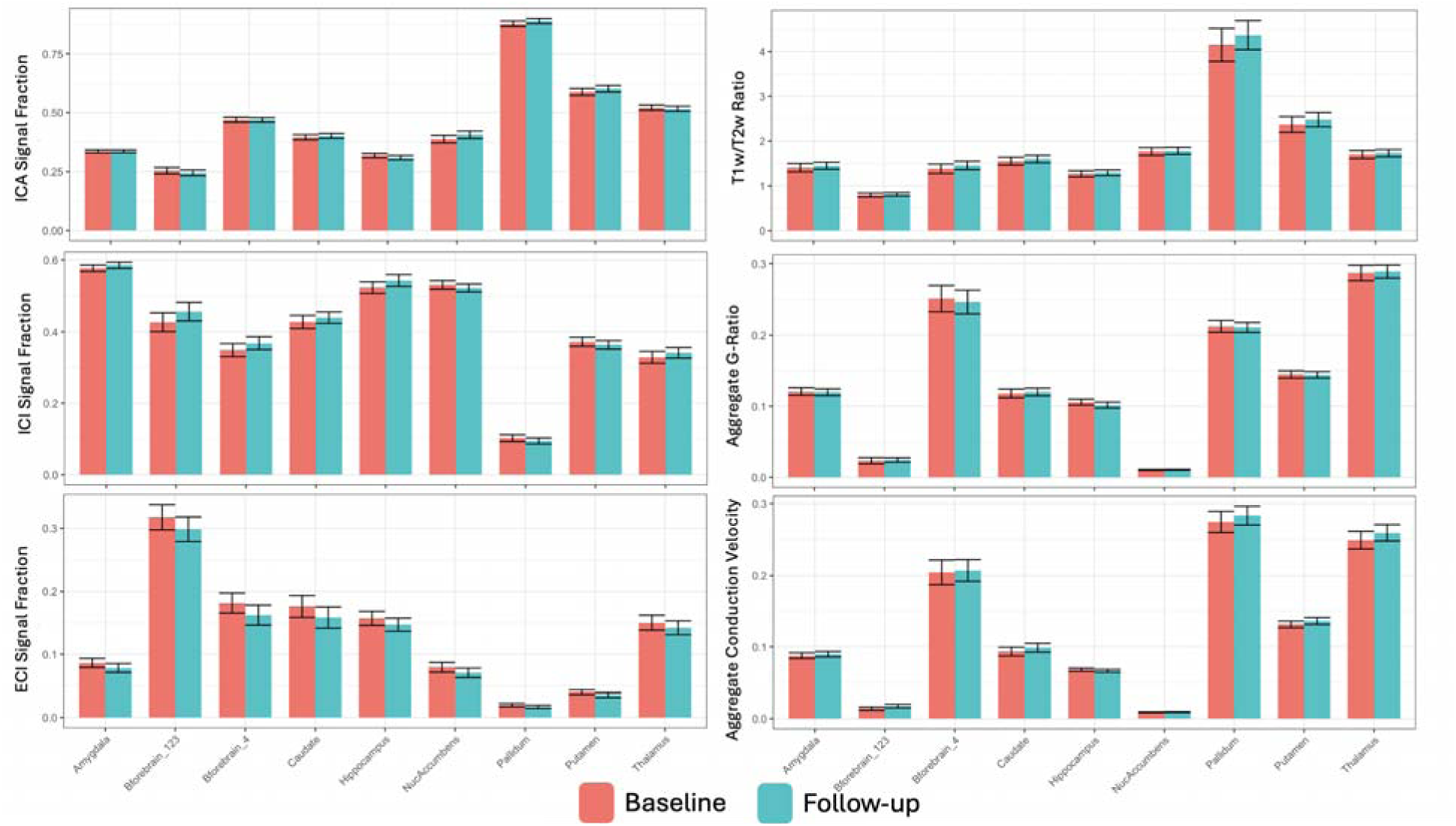
Charts displaying baseline and follow-up means and standard deviations for each microstructural metric in each ROI examined. Despite the focus on similar subcortical gray matter structures there was a wide degree of variation in absolute microstructural values in the ROIs examined. No change from Baseline to Follow-up was significant after multiple comparison correction. All values displayed in these charts is listed in Supplemental Table 1. Abbreviations: ECI, extracellular isotropic; ICI, intracellular isotropic; ICA intracellular anisotropic ICA.

**Figure 4:**
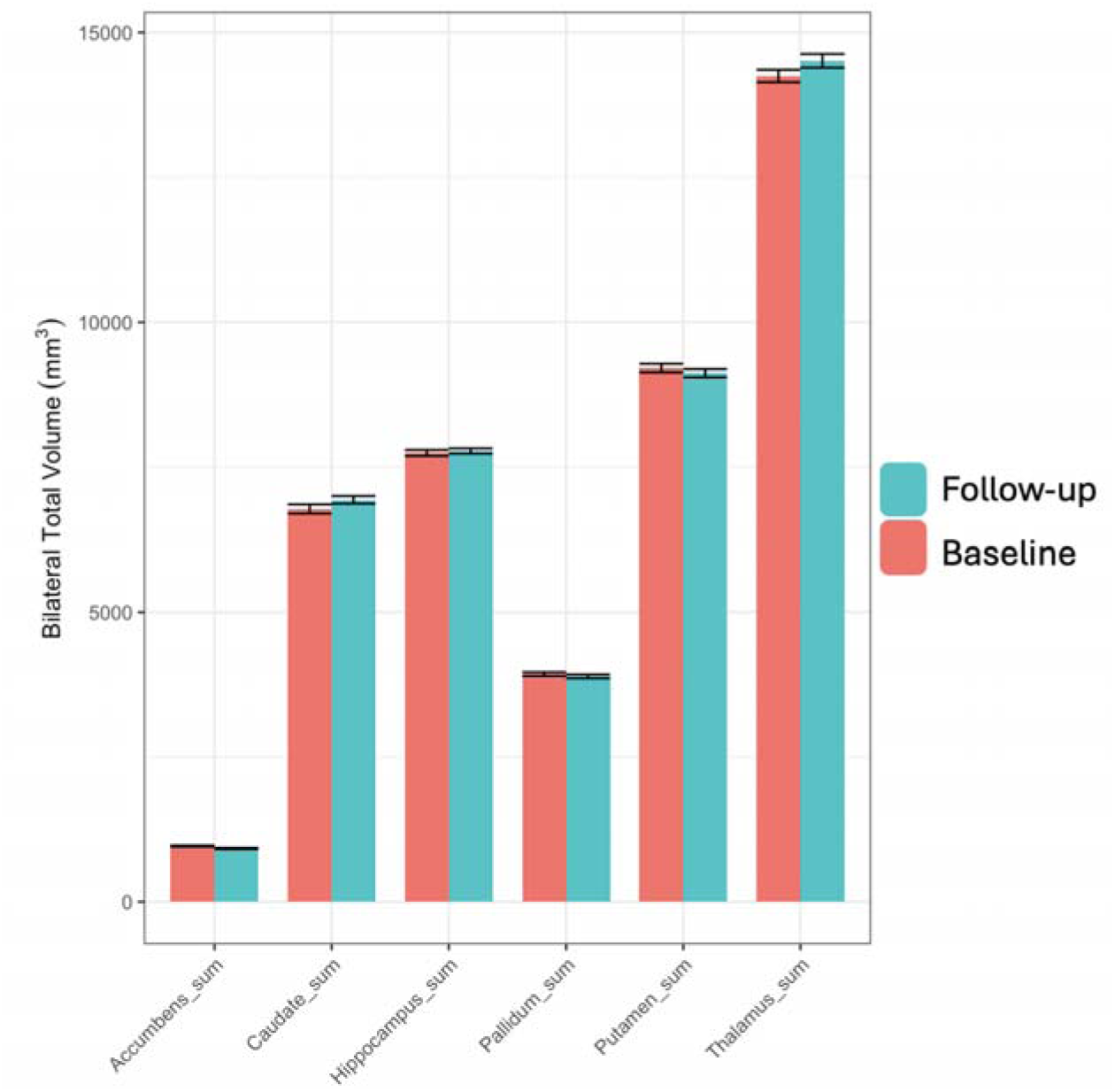
Charts displaying the baseline and follow-up volumes (group-means and standard deviations) for each ROI volumetric measurement available from the Freesurfer recon-all analysis pipeline. There was a split in the direction of volumetric changes, with the thalamus, hippocampus, and caudate significantly increasing and the putamen and pallidum significantly decreasing between baseline and follow-up. There was a nonsignificant decrease in the volume of the accumbens between baseline and follow-up. Associations of MR Metrics With Longitudinal Changes in Motor Symptoms, Cognitive Symptoms, and Latent Fitness

Main effects of volumes and microstructural metrics in each of the nine subcortical ROIs on motor symptoms, cognitive symptoms, and fitness can be found in Supplemental Tables 2-7 (2: MOCA, 3: Executive Function Z-Score, 4: Attention Z-Score, 5: Language Z-Score, 6: Memory-Visuospatial Z-Score, 7: Latent Fitness controlled for baseline fitness). Table 2 list statistical results for regional associations between baseline MR metrics and longitudinal changes in symptom and fitness scores that remained significant after FDR correction. In the caudate, changes in Executive Function, Attention, and Memory and Visuospatial Z-Scores (all *p*<0.05) varied significantly with ICI. ICI had a sparingly significant relationship with change in the Language Z-Score (*p*=0.053). The volume of the caudate was also significantly associated with change in the Attention Z-Score (F_1_=10.06, *p*<0.01). In the putamen, changes in Executive Function and Memory-Visuospatial Z-Scores varied with ICI (both *p*<0.05). In the nucleus accumbens, changes in Executive Function and Memory-Visuospatial Z-Scores varied significantly with ICI (both *p*<0.05) and change in the Language Z-Score varied sparingly significantly with ICI (*p*=0.053). ICI of the thalamus varied significantly with change in the Executive Function Z-Score (*p*<0.05). In Ch4, ICI varied with change in Latent Fitness (*p*<0.01) and *ICA* varied with change in MOCA (*p*<0.01). Volumetric associations with motor symptoms, cognitive symptoms, and fitness can be found in Supplemental Table 8, the bilateral volume of the caudate was significantly associated with Attention (p<0.01). No other significant associations were found between MR metrics and symptom or fitness scores in any ROI examined.

**Table 2:** Associations of regional MR metrics at baseline with longitudinal changes in motor and cognitive scores and fitness. For each ROI-symptom domain combination, MR metrics are listed that had a significant relationship with the symptom or fitness score (p-values for linear mixed model, FDR-corrected). ns: no significant findings. The ICI metric (reflecting mainly diffusion within neuronal and astrocytic perikarya) in three striatal nuceli (caudate, putamen, accumbens) and the thalamus predicted change in multiple cognitive domains. Caudate volume additionally predicted change in attention. In Ch4, ICA (reflecting mainly diffusion within axons and neurites) predicted change in global cognition (MOCA) and ICI predicted change in fitness. No other volume or microstructure-symptom associations were significant after multiple comparison correction.

| **ROI** | **MOCA** | **Executive Function** | **Attention** | **Language** | **Memory & Visuospatial** | **Latent Fitness** |
| --- | --- | --- | --- | --- | --- | --- |
| *Amygdala* | NA | NA | NA | NA | NA | NA |
| *Cholinergic Basal Forebrain Area 1,2,3* | NA | NA | NA | NA | NA | NA |
| *Cholinergic Basal Forebrain Area 4* | ICA p<0.01 | NA | NA | NA | NA | ICI p<0.01 |
| *Caudate* | NA | ICI p<0.05 | ICI p<0.05 | ICI p=0.053 | ICI p<0.05 | NA |
| *Hippocampus* | NA | NA | NA | NA | NA | NA |
| *Nucleus Accumbens* | NA | ICI p<0.05 | NA | ICI p=0.053 | ICI p<0.05 | NA |
| *Pallidum* | NA | NA | NA | NA | NA | NA |
| *Putamen* | NA | ICI p<0.05 | NA | NA | ICI p<0.05 | NA |
| *Thalamus* | NA | ICI p<0.05 | NA | NA | NA | NA |

### Diffusion Tractography

The number of tracts connecting the CH4 with known afferent subcortical structures was examined in models identical to the previous tests (Fig. 5). The connectivity between the CH4 and thalamus was significantly associated with baseline motor skill fitness (F=5.176, p<0.05) and was nearly significantly associated with follow-up fitness (F=4.33, p=0.059, n.s.) and with the MOCA (F=4.03, p=0.066, n.s.). This matched microstructure results for the CH4 generally. The connectivity between the CH4 and the caudate, putamen, and amygdala was not significant for any of the behavioral metrics tested in this study. The full list of all statistical tests between connectivity and behavioral metrics is available in Supplemental Table 8.

**Figure 5:**
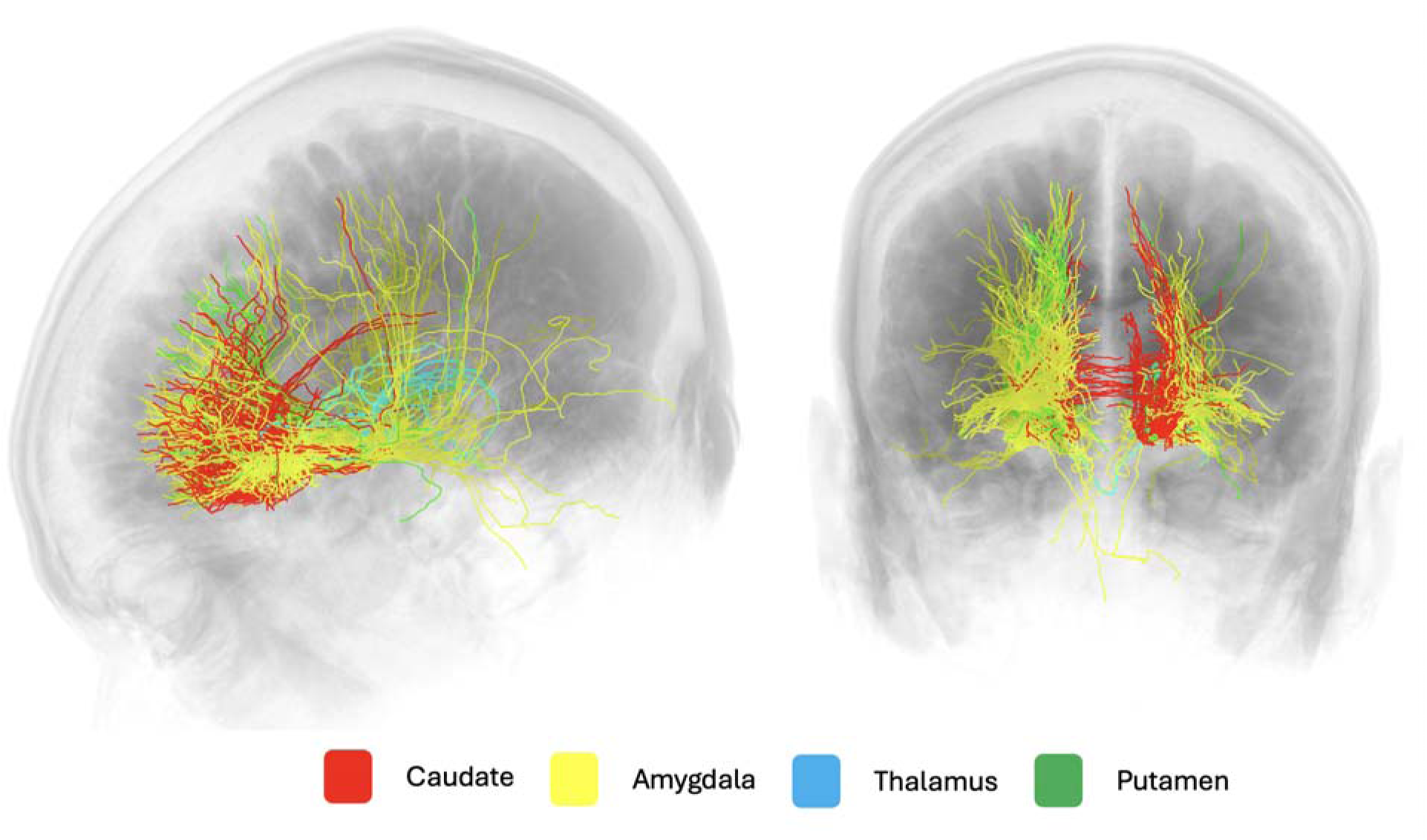
example from a single subject showing connections between CH4 and the caudate, amygdala, thalamus, and putamen. All regional masks were applied bilaterally throughout the cohort. Tracts that crossed through both CH4 and the relevant ROI were counted to capture all available connections from the probabilistic tracking method.

## Discussion

This study examines a longitudinal cohort of PD patients using advanced diffusion microstructure analysis to evaluate several regions of the subcortical gray matter and basal ganglia for associations with clinically relevant cognitive measures and level of motor skill fitness. Associations wer found between executive, attention, language, cognition, and motor skill fitness scores and intracellular isotropic signal across 5 subcortical and basal ganglia ROIs (caudate, putamen, thalamus, nucleus accumbens, CH4). Executive function and the cognitive composite score demonstrated significant correlation with the intracellular isotropic signal across all these regions, excluding the basal forebrain area, CH4. Intracellular anisotropic signal was related to MOCA score in CH4. Volumetrically, there was a significant change between subject baseline and follow-up in 5 ROIs, however none of these changes was correlated to measures of cognitive function or motor skill fitness. Structural connectivity between the CH4 and thalamus were found to be significantly associated with motor skill fitness.

Taken together, these findings suggest that the ICI metric in the striatum and thalamus fairly prognosticates evolution in specific cognitive domains in PD. Global cognitive status and exercise-mediated recovery of cognitive function may be mediated in part thru axonal projections, including to the thalamus.

Although 6 separate microstructure metrics were assayed, nearly all significant associations with cognitive symptoms and fitness involved a single metric, the ICI. Crucially, as the signal fractions (ICI, ICA, ECI) are summed to unity, the changes observed in this study are relative to the signal from each of the other compartments, and are not necessarily indicative of change in the overall signal. Observing the longitudinal changes over time in this cohort, the increase in intracellular isotropic signal appears to occur with a decrease in extracellular free water, with intracellular anisotropic signal unchanged. The increase in extracellular free water is typically thought to arise from cellular atrophy and degeneration, as cellular bodies are lost the resulting extracellular space is filled with freely diffusing fluid^11,84–86^. An increase in intracellular isotropic signal however, is thought to involve increased neuronal cell somas or glial structures, and has previously been implicated in events such as pubertal development in adolescents^53^. In both the intracellular signal fraction change observed in this study, and the increase observed in puberty, it is proposed that conformational changes in glial cells drive the observed microstructural change. Pubertal development is characterized by increased myelination throughout the cortex, which is performed by oligodendrocytes, and it is likely that observed intracellular isotropic signal increases reflect oligodendrocyte proliferation and growth. In the current study, during the opposite end of the lifespan, the pathology of PD may be inducing reaction from various glial cells. Prior work in rodent models suggests that astrocyte activation and conformational change occurs during PD^17^. This has been suggested to be mediated through exercise in PD, and in this study we observe increased intracellular isotropic signal associated with motor skill fitness, potentially from these activated astrocytes. This was observed to especially be the case in the CH4, a critical region associated with PD progression and symptomology^5–7^.

Consistent with a glial origin of our present findings is the lack of significant symptom associations with or longitudinal changes in the four microstructural metrics linked to axonal structure and function. T1w/T2w ratio measures myelin integrity, and ICA, aggregate g-ratio, and aggregate conduction velocity all characterize axonal properties. While the ROIs examined in this study were predominantly gray matter structures, like subcortical nuclei and cortex generally, they are still typically innervated by or act as the source of axons and, thus may exhibit axonal effects on microstructural indices^52^. The effect observed in this report for Ch4-thalamus structural connectivity may reflect an influence of axonal properties in the intervening white matter on attention and fitness in PD.

Interestingly, several subcortical structures exhibited longitudinal volume changes in the absence of microstructural effects between baseline and follow-up. Though microstructural changes have previously been shown to predate volumetric changes, the primary tissue compartment involved in these changes has typically been the extracellular free water compartment^85^. Free water in the substantia nigra has also featured large in previous microstructure work in PD and has been proposed as a measure of the functional status of surviving dopaminergic nerve terminals^11^. It is possible that the timespan between baseline and follow-up was too short for such effects to manifest, as no microstructural changes were significant between baseline and follow-up in the current study. The overall small number of participants who were able to complete the extensive battery of all imaging, cognitive, and fitness metrics may have contributed to less than expected longitudinal results.

Prior work tying altered fractional anisotropy to PD symptomology of various kinds supports the notion of altered microstructural properties in the brain in PD. A recent meta-analysis found increased mean diffusivity was particularly sensitive to clinical PD parameters, similar to the ICI findings of our study^87^. Compared to mean diffusivity, ICI is more specific than mean diffusivity to isotropic signal within cellular bodies. Mean diffusivity additionally includes the presence of the extracellular isotropic compartment. Thus, increased mean diffusivity may result from either increased cellularity or increased free water, the results of this study suggest that increased ICI is due to cellular change and not increased extracellular water. Some studies report an unexpected increase in fractional anisotropy in specific white matter tracts in PD. The positive associations observed in this study between Ch4-thalamus structural connectivity and attention and Ch4 ICA and MOCA score are consistent with findings of this kind.

This study utilizes a number of advanced diffusion microstructure metrics to evaluate a longitudinal cohort of PD patients. By finding widespread associations between an isotropic intracellular signal and cognitive and fitness metrics, this study suggests that increased cellularity, potentially from astrocytic glial cells, may support reduced symptoms of decline during PD as well as increased level of motor skill fitness.

## Data Availability

All data produced in the present study are available upon reasonable request to the authors.

